# Cost-effective and scalable clonal hematopoiesis assay provides insight into clonal dynamics

**DOI:** 10.1101/2023.11.08.23298270

**Authors:** Taralynn Mack, Caitlyn Vlasschaert, Kelly von Beck, Alexander J. Silver, J. Brett Heimlich, Hannah Poisner, Henry Robert Condon, Jessica Ulloa, Andrew L. Sochacki, Travis P. Spaulding, Ashwin Kishtagari, Cosmin A. Bejan, Yaomin Xu, Michael R. Savona, Angela Jones, Alexander Bick

**Author notes:** **Correspondence to:** Alexander G. Bick, 550 Robinson Research Building, Vanderbilt University Medical Center, 2200 Pierce Ave, Nashville, TN 37232.

## Abstract

Clonal hematopoiesis of indeterminate potential (CHIP) is a common age-related phenomenon that occurs when hematopoietic stem cells acquire mutations in a select set of genes commonly mutated in myeloid neoplasia which then expand clonally. Current sequencing assays to detect CHIP are not optimized for the detection of these variants and can be cost-prohibitive when applied to large cohorts or serial sequencing. Here, we present and validate a CHIP targeted sequencing assay that is affordable (∼$8/sample), accurate and highly scalable. To demonstrate the utility of this assay, we detected CHIP in a cohort of 456 individuals with DNA collected at multiple timepoints in the Vanderbilt BioVU biobank and quantified clonal expansion rates over time. A total of 101 individuals with CHIP were identified, and individual-level clonal expansion rate was calculated using the variant allele fraction (VAF) at both timepoints. Differences in clonal expansion rate by driver gene were observed, but there was also significant individual-level heterogeneity, emphasizing the multifactorial nature of clonal expansion. We further describe the mutation co-occurrence and clonal competition between multiple driver mutations.

## Introduction

Although our DNA is frequently thought of as fixed at conception, recent studies have described how our genome changes as a natural consequence of aging. Our telomeres shorten, mitochondrial DNA evolves in copy number and sequence, and our nuclear genome accumulates mutations.^1–3^ Hematopoietic stem cells (HSCs) –the self-renewing progenitors of all circulating blood cells– accumulate mutations as they divide. Individual HSCs are estimated to acquire 200 mutations per decade genome-wide, with one mutation per decade occurring within an exonic region.^4–6^ The vast majority of the time, these somatic mutations confer little to no advantage to the cell, but infrequently fitness-increasing “driver” mutations confer a proliferative advantage.

Clonal hematopoiesis of indeterminate potential (CHIP) is an age-related condition that is characterized by the acquisition of a somatic driver mutation in an HSC that leads to an expanded blood cell lineage, termed a clone.^7,8^ The condition increases in prevalence with age, and occurs in >10% of individuals over the age of 70.^9,10^ CHIP confers an increased risk of hematologic cancer,^2^ cardiovascular disease,^11^ COPD,^12^ liver disease,^13^ kidney disease,^14,15^ osteoporosis,^16^ and overall mortality.^11^ The minimum variant allele fraction (VAF) used as a threshold to define CHIP is 2% (corresponding to 4% of circulating diploid cells).

CHIP-associated morbidity generally increases proportional to the VAF.^2,11,17,18^ Therefore, the ability to identify individuals who will likely have a faster clonal growth rate, leading to a higher VAF, is important to prevent unfavorable CHIP outcomes. However, quantifying clonal expansion rate is difficult due to a current scarcity of multi-timepoint data precluding observation of these behaviors. In order to understand the factors that impact clonal behavior over time, identifying CHIP accurately at low cost and at scale is essential.

CHIP can be identified in peripheral blood through a variety of sequencing approaches including whole-genome sequencing,^9,18^ whole-exome sequencing,^2^ or through targeted sequencing approaches.^19,20^ Whole genome/exome sequencing are expensive and have limited sequencing depth, making these approaches an unsuitable solution to the large-scale detection of CHIP clones. Targeted amplicon-based approaches are less expensive (∼$30-50/sample) but are limited in size and have technical challenges precluding robust detection of specific common CHIP loci such as *ASXL1* or *SRSF2* where the high GC content and repetitive sequences makes amplicon design difficult. Conversely, hybrid capture based approaches can overcome some of the shortcomings of amplicon design, but historically have been significantly more expensive ($50-150/sample).

To address this sequencing limitation, we developed a highly cost-effective hybrid capture based targeted gene panel that encompasses 95% of CHIP found in the general population^9^ by sequencing 22 CHIP genes at high depth of coverage (∼2,000x, **Figure 1**). This method increases CHIP detection accuracy and allows even small clones to be detected in the blood.

**Figure 1:**
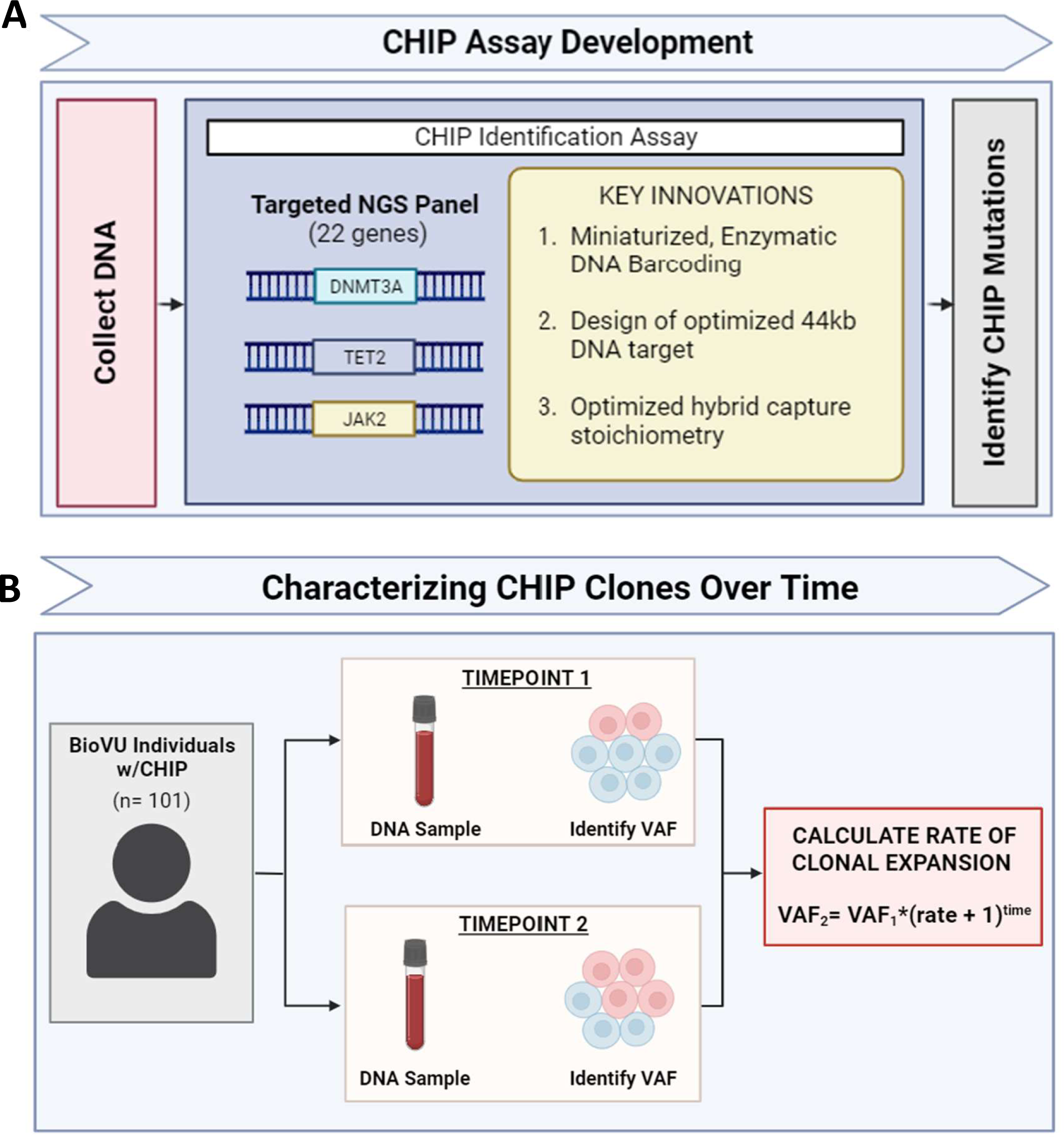
Study Design. a) Schematic representation of CHIP Identification Assay. DNA is extracted from the blood of an individual and tested for CHIP using the low-cost, scalable targeted assay that tests 22 specific genes and positions. CHIP mutations are identified and their variant allele frequency is estimated. b) Schematic representation of characterizing CHIP clones over time in BioVU. The CHIP assay was applied to a cohort of individuals in Vanderbilt BioVU with multiple blood samples over time. CHIP was identified in one or both timepoints and VAF was estimated, allowing for the estimation of clonal growth rate over time and characterization of clonal behavior.

As a demonstration of the potential utility of this tool, we applied this assay to a multi-time point blood draw cohort of individuals from Vanderbilt BioVU (n =456) and identified 101 individuals with CHIP. We then quantified clonal expansion rate on a per-individual basis, which allowed us to identify the driver genes with faster or slower average rates of expansion. In individuals with multiple CHIP driver genes, we observed instances of opposing clonal behavior, possibly indicating clonal competition. Together, this method and the subsequent findings help to better characterize CHIP clonal behavior over time and identify specific CHIP mutations at the highest risk of rapid expansion.

## Methods

### Vanderbilt BioVU Samples

The Vanderbilt University Biobank (BioVU) is a de-identified biorepository with ∼300,000 DNA samples extracted from leftover peripheral blood samples used for routine clinical care. All BioVU participants have provided informed consent. The VUMC Institutional Review Board oversees BioVU and approved this project (IRB#201783). Additional DNA samples were collected as the original DNA sample was depleted and as a result ∼30,000 BioVU participants have blood samples taken at multiple timepoints over up to 15 years.^21^ The 456 BioVU participants sequenced for this study, were selected based on signals of somatic mosaicism from the BioVU Illumina MEGA genotyping array to maximize the number of individuals with longitudinal CHIP trajectories (**Supplementary Figure 1, Supplemental Table 1)**. DNA extraction for BioVU leveraged the AutoGen flexstar instruments and was quantified with nanodrop and ThermoFisher Quant-iT picogreen assay.

While all 456 individuals were sequenced on the CHIP assay at multiple timepoints, the 101 individuals used in downstream analyses were a subset of those individuals with at least one CHIP mutation (VAF > 2%) and without a blood cancer diagnosis (**Supplemental Figure 1**). Patient charts were reviewed to determine blood cancer status and to extract other relevant phenotypic and demographic data.

### Development of a <$10 CHIP Sequencing Assay

We developed a novel, highly scalable and highly cost-effective method to identify individuals with CHIP (**Figure 1a**). In order to prioritize which CHIP genes to include in the panel, we analyzed the distribution of genes in 4,229 individuals with CHIP from the TOPMed cohort. As has been previously described, CHIP has a skewed distribution: >60% of CHIP mutations are found in just two genes (*DNMT3A* and *TET2*). >95% of CHIP mutations are present in the 24 most common genes (**Supplemental Figure 2**).^9^ Building on this analysis we identified a specific set of DNA regions that encompass just ∼44kb of DNA but encompass >95% of all CHIP observed in the general population. The genes include: *ASXL1, ASXL2, BRCC3, CBL, DNMT3A, ETNK1, GNAS, GNB1, IDH1, IDH2, JAK2, KIT, KRAS, MPL, NRAS, PPM1D, SETBP1, SF3B1, SRSF2, TET2, TP53, and U2AF1.* (**Supplemental Table 2**). The cost of sequencing such a small region of DNA, even at >1000x coverage is negligible (<$0.30) on an Illumina NovaSeq 6000 with a S4 flow cell). We designed hybrid capture oligonucleotide probes to selectively amplify this set of genomic intervals (Twist Bioscience).

Next, we sought to develop a highly scalable and cost-efficient method for selectively sequencing these CHIP DNA intervals. We began by developing a scalable genomic library preparation method that is compatible with the hybrid capture system to enable efficient processing of genomic DNA from multiple individuals at the same time through DNA barcoding. Our approach leverages the Twist Library Preparation Enzymatic Fragmentation Kit 2.0 (Twist Bioscience, #104207). Comparison of standard mechanical fragmentation and this enzymatic library preparation demonstrated highly concordant results. We further optimized the manufacturer’s protocol through 5x miniaturization of the amount of input reagents for each unit of DNA which significantly reduced the cost.

The hybrid capture technology was optimized to selectively enrich CHIP regions of interest from the pooled DNA libraries. We found that with the custom designed hybrid oligonucleotide probes we could perform 12 times the number of hybridization reactions as the manufacturer’s protocol (96 DNA plex samples for each reaction of hybrid capture probes). We then established this entire pipeline on a standard liquid handling robot platform (Biomek i7) to enable preparation and sequencing (on a single liquid handling robot) of up to 5,000 samples/week.

Utilizing this approach, we sequenced the CHIP enriched DNA libraries on standard next-generation sequencing chemistry (Illumina NovaSeq 6000). We found that these libraries were compatible with other standard barcoded libraries such that we could spike in 1-5% of the CHIP assay to the total pool flow cell pool which typically would contain whole genome sequencing samples. This enables sequencing smaller batches of several hundred samples at a time, rather than waiting for the tens of thousands of samples it would take to fill entire flow cells with just the CHIP assay. At the same time, we are able to also take advantage of the greater per-base cost efficiency afforded by the larger flow cells.

Putative somatic mutations are identified in the aligned sequencing reads using the Mutect2-GATK package, and filtering is applied to identify variants that met previously described criteria for CHIP.^18^ Variants with total low read depth (<100), low variant allele read depth (<3) and/or variant allele fraction below the threshold for CHIP (<2%) were removed from the dataset.

### Calculating Rate of Clonal Growth

Clonal fitness was calculated on a per-individual basis using an exponential growth rate model:

**VAF_2_= VAF_1_* (CF +1)^time^**

Based on this exponential model, clonal fitness was calculated as such:

**CF= ((VAF_2_/VAF_1_) ^(1/(Time))^) −1**

For individuals with CHIP detected >2% at only one timepoint, the missing VAF was set to 0.001 to reflect the sequencing detection limit of the assay.

### Classifying Clonal Trajectories

We categorized each CHIP clone into a “expansion”, “reduction”, or “stagnant” category. To do this, we took the bottom 10% of clones based on the absolute value of their growth rate and categorized them as “stagnant”. This corresponded to less than 2% annual growth rate. For the remaining 90% of clones, clones with a positive growth rate were grouped into the “expansion” category and clones with a negative growth rate were grouped into the “reduction” category.

Individuals with two driver mutations were also grouped into “distinct clone” and “sub-clone” categories (**Supplemental Figure 3**). We z-scored the “expansion”, “stagnant”, and “reduction” clone groups separately, producing an individual z-score for each clone. Individuals with <0.6 difference in z-scores between their clones growing in the same direction were assigned to the sub-clone category. Individuals with clones growing in opposite directions or whose clones had >=0.6 difference in z-scores were categorized as distinct clones.

### Association between clonal expansion rate and participant characteristics

Using a linear/logistic regression model, we tested associations between CHIP expansion rate and age, gender, self-reported race, ethnicity, BMI, and height individually. CHIP driver gene was used as a covariate in each model.

## Results

### CHIP Assay Validation

To quantify the limit of detection for our method we performed a limiting dilution experiment where a DNA sample with known genotype was combined at serial fixed ratios with a second sample of known genotype (**Supplemental Figure 4**). We utilized these ratios to identify the expected allele fraction for a given variant. We find that our method robustly detects variants present in >1% of DNA. Furthermore, we find that beneath this 1% threshold, we continue to detect variants down to ∼0.3% allele fraction, but with less accuracy for the estimated allele fraction.

For further validation of our assay in comparison to a CLIA reference laboratory test gold standard (Vanderbilt Molecular Diagnostics Laboratory, Myeloid Next Generation Sequencing panel), we considered a set of clinical patient samples from our Vanderbilt CHIP clinic. 41 out of 41 patients with CHIP from the clinical assay were identified as having CHIP on our assay.

### CHIP prevalence and characteristics in Vanderbilt BioVU cohort

We applied our CHIP assay to a set of 456 individuals with two or more DNA samples available from different timepoints in the Vanderbilt University Biobank (BioVU) (**Figure 1b**). These 456 individuals were selected from a pool of ∼30,000 individuals in BioVU with multiple blood draws, which is a subset of the general Vanderbilt BioVU biobank (n∼300,000) (**Supplementary Figure 1**). These individuals were selected based on suggestion from the BioVU MEGA genotyping array intensity data that they were likely to have CHIP in *DNMT3A*, *TET2* or *JAK2.* These individuals were of increased age (Timepoint 1: mean: 52.5 years, SD: 19 years; Timepoint 2: mean: 57 years, SD: 19 years) and were 60% female and 81% Caucasian (**Supplemental Table 1**). Samples were sequenced to an average depth of 1,725x. Of these individuals, 173 (34%) had CH driver mutations with VAF >2% at at least one time point. 72 individuals were removed based on a blood cancer diagnosis that either predated the DNA collection or occurred within 6 months of the DNA collection, suggesting they had blood cancer rather than CHIP.

Our final cohort included 101 individuals with CHIP and 145 driver mutations. 66% of individuals had a single CHIP mutation, 26% had two mutations, and 8% of individuals had 3 or more CHIP mutations (**Figure 2a**). The most common CHIP driver mutation was *DNMT3A,* followed by *TET2* (**Figure 2a**), which is consistent with previously reported driver gene distributions.^9^ 40/73 (55%) mutations in *DNMT3A* were at recurring hotspots R882C/R882H, likely because these mutations were directly genotyped on the MEGA assay.

**Figure 2:**
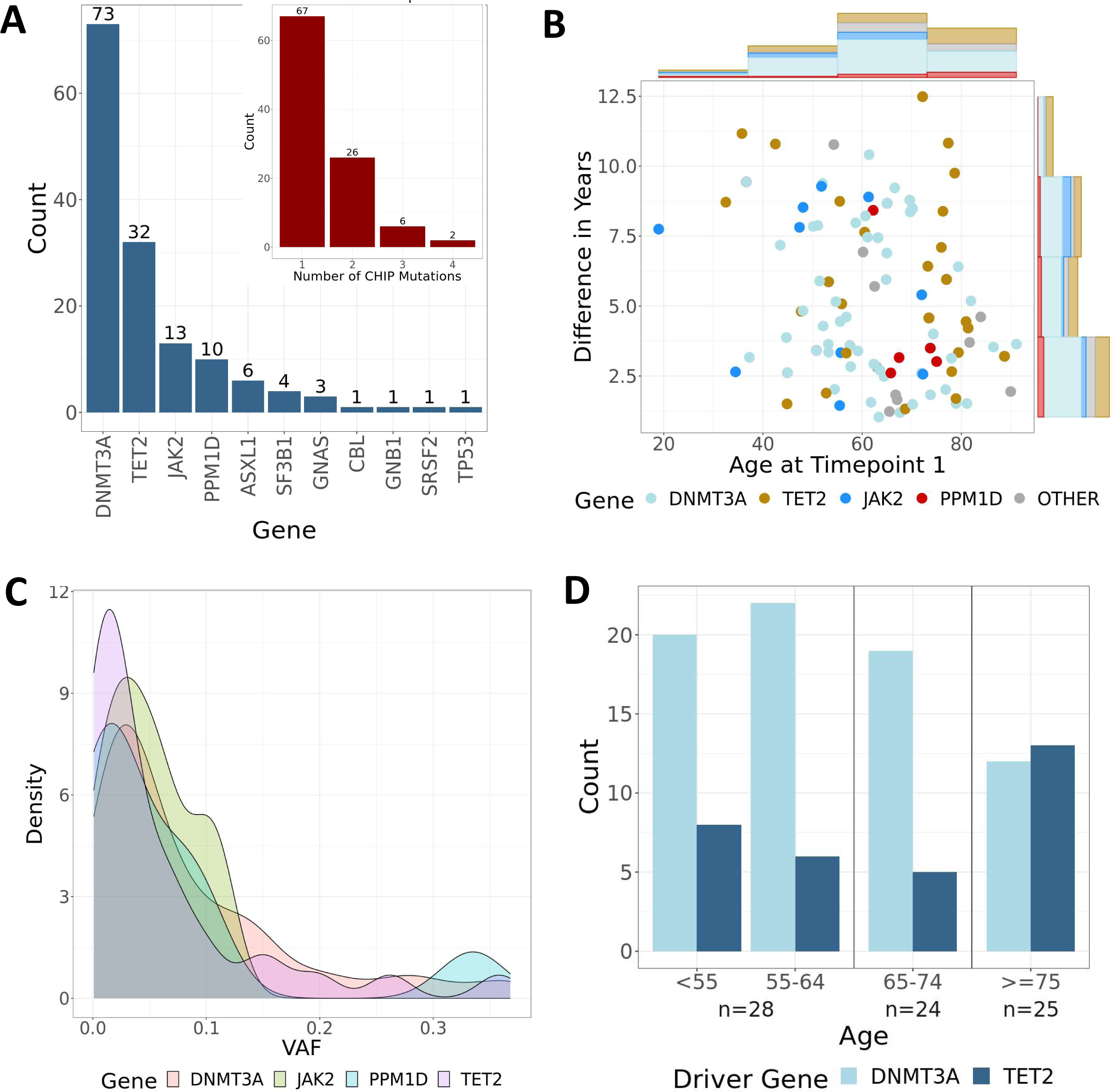
CHIP detected in the BioVU cohort. a) Bar plots showing the counts of mutations present in the cohort per driver gene and the counts of individuals with different numbers of CHIP driver mutations. *DNMT3A* and *TET2* are the most common driver genes and the vast majority of individuals have one CHIP driver mutation. b) Plot showing the distribution of age at timepoint 1 as well as the difference in years between the time points for each individual in the cohort. Histograms along the axes reflect total counts as well as the driver gene distribution. d) Density plots showing the distribution of VAF at timepoint 1 across the most common driver genes. There was no significant difference between driver genes. d) Bar plot showing the distribution of *TET2* and *DNMT3A* driver mutations across age categories.

Within the subset of individuals with CHIP, the average age at timepoint 1 was 65 years (range: 18-91 years), and the average age at timepoint 2 was 70 years (range: 26-94 years) (**Supplemental Table 1**). The average time between the two timepoints was 5.4 years (range: 1-12.5 years) (**Figure 2b**). Average VAF at timepoint 1 was 8%, and 11.5% at timepoint 2. Average VAF did not significantly differ between CHIP driver genes (**Figure 2c**). For individuals 75 and older, *TET2* surpassed *DNMT3A* as the most common CHIP driver mutation (**Figure 2d**).

### Clonal expansion rate across driver genes

Since different CHIP driver genes can confer different risks and mechanisms of action,^9,10,22^ we compared expansion rates between different driver genes. Across mutations with at least 10 individuals, *JAK2* clones showed the fastest rate of expansion on average (109% growth/year) and *DNMT3A* clones showed the slowest rate (8% growth/year) (**Figure 3a**). To eliminate the possibility of expansion rates being skewed by the presence of multiple clones in an individual, we performed a sensitivity analysis that restricted the individuals to only those with a single CHIP driver mutation. Although this reduced our sample size by 30%, *JAK2* clones remained the fastest growing (178% growth/year), and *DNMT3A* clones remained the slowest growing (5% growth/year).

**Figure 3:**
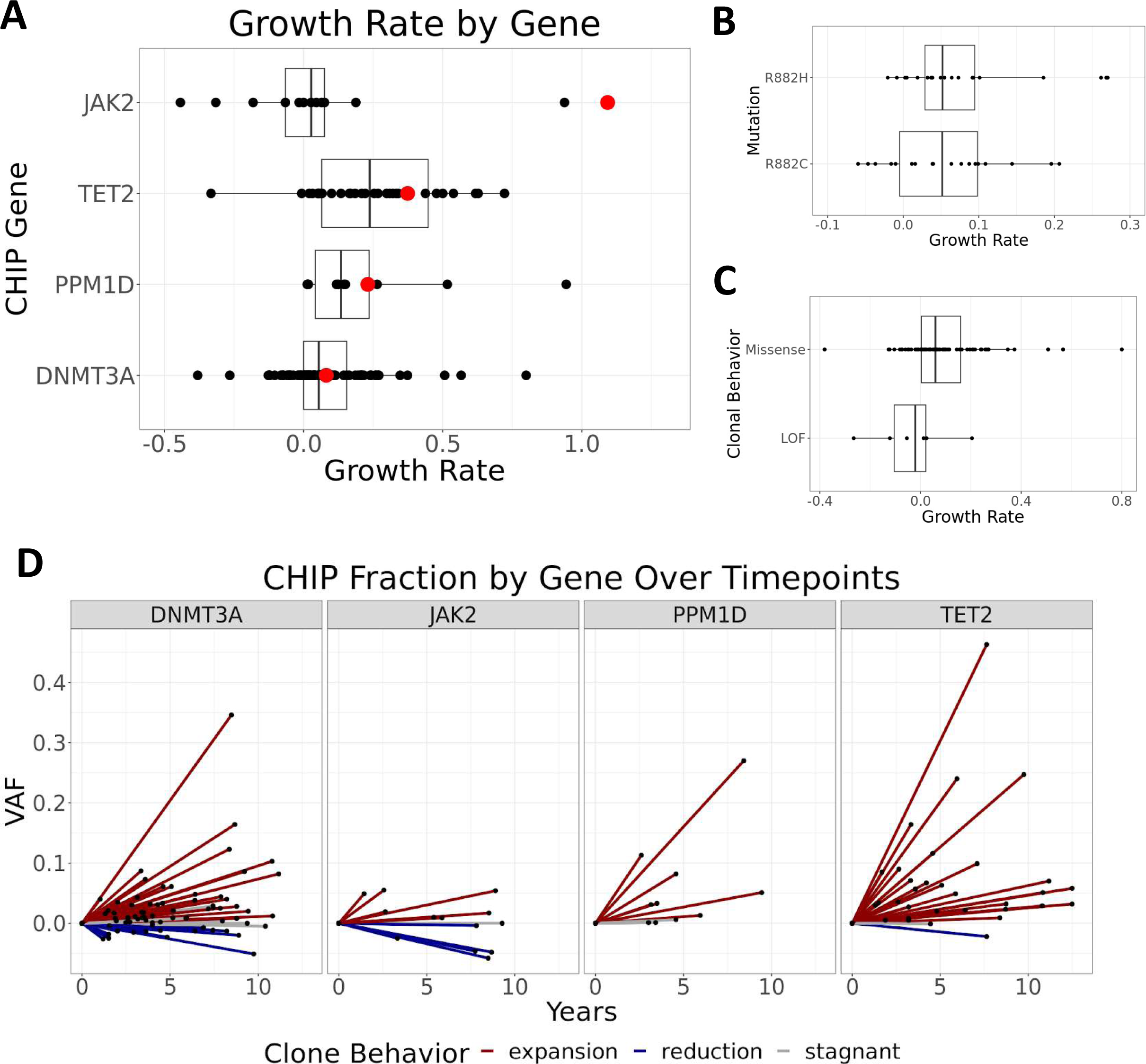
CHIP clonal behavior by driver gene. a) Average growth rate across driver gene for all individuals with CHIP mutations. Each point represents one individual. Genes are arranged in descending order on the y-axis based on mean growth rate, and the x-axis is limited and excludes values greater than 1.2. Average growth rate is marked by a red dot. b) CHIP clone trajectories are shown by change in VAF over time. Each line represents one clone while the length of the line represents years in between timepoints and the slope of the line represents net change in VAF over time. Lines are colored by clone behavior category and are stratified by CHIP driver gene.

Of the 73 *DNMT3A* mutations, 21 of them were R882H and 19 were R882C, which are known hotspots. No significant difference in expansion rate was observed between these two different hotspot mutations (p=0.63) (**Figure 3b**). Stratifying *DNMT3A* mutations into missense *versus* loss of function identified a non-significant trend towards increased growth amongst individuals with missense mutations (p=0.08) (**Figure 3c**).

### CHIP clonal dynamics

Based on the change in VAF between the timepoints, we classified each clone into qualitative categories: “expansion”, “reduction”, and “stagnant”. 78% of clones grew over time, showing that once an individual acquires CHIP that clone tends to keep growing. However, instances of clonal shrinkage were observed across driver mutations (**Figure 3d**), even when restricting to individuals with only one driver mutation. The clones that shrank comprised 40% of *ASXL1* clones, 26% of *DNMT3A* clones, 46% of *JAK2* clones, 6% of *TET2* clones, and 13% of *PPM1D* clones. *ASXL1* clones showed the fastest rate of shrinkage on average (13% shrinkage/year) and *TET2* clones showed the slowest rate of shrinkage (0.6% shrinkage/year).

Within individuals with more than one driver mutation, it is possible that the mutations are in distinct cell populations (distinct clones) or that the two mutations co-occur in the same cell populations (sub-clones) (**Figure 4a**). Using clone trajectory, we identified which of these scenarios was more likely for individuals with two mutations (see Methods) and classified each person into one of the two groups (distinct clones vs. sub-clones). Out of the 26 individuals with two mutations, 12 individuals had distinct clones and 14 had sub-clones (**Figure 4b**). When we compared the average growth rate of each individual’s fastest growing clone by group, there was no significant difference between the distinct and sub-clone groups (p=0.20). When looking at the combinations of driver genes, *DNMT3A* mutations are most likely to remain stagnant or shrink in the presence of an additional driver mutation compared to other drivers (**Figure 4c**).

**Figure 4:**
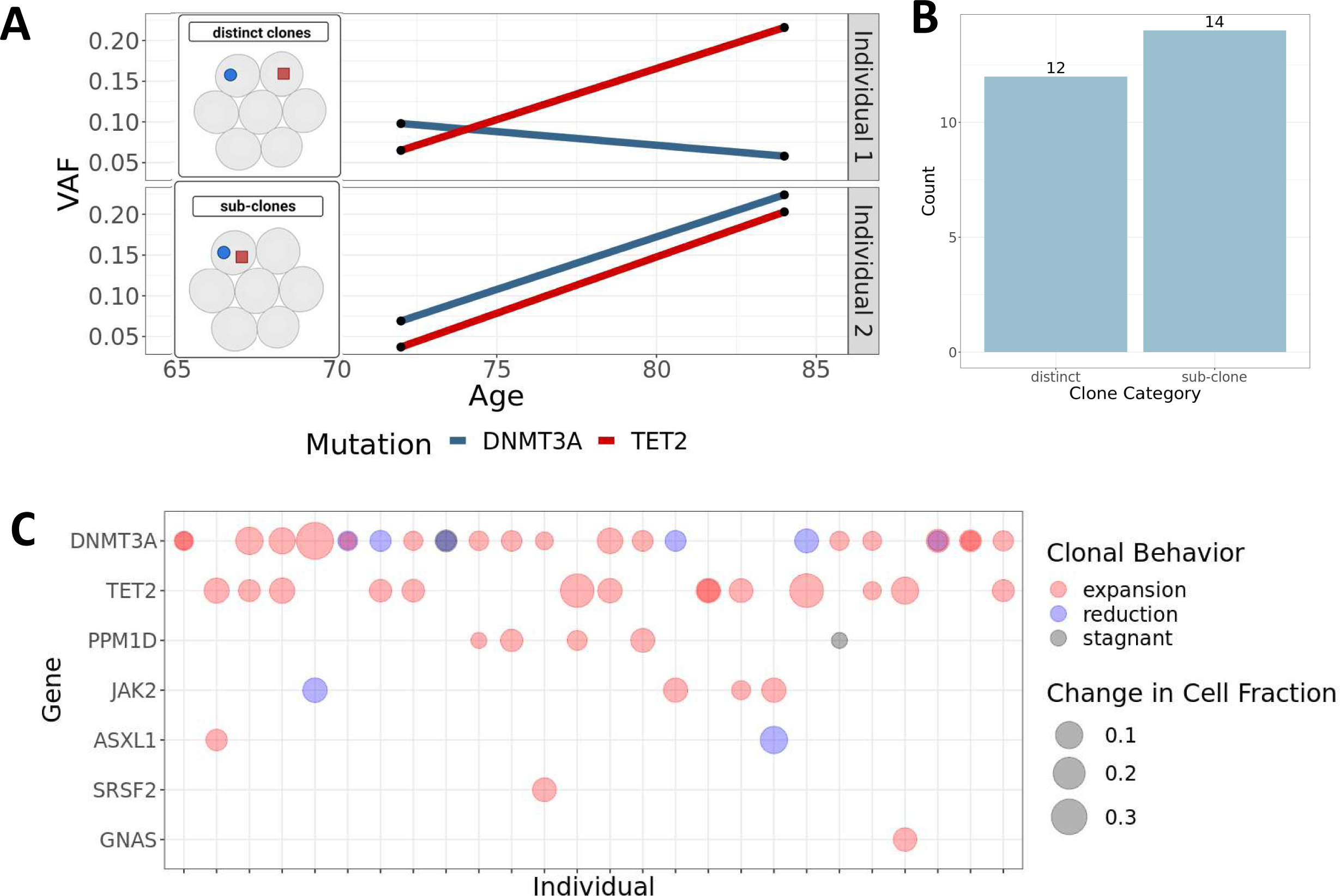
Clonal trajectories over time in individuals with more than one driver mutation. a) For individuals with more than one driver mutation, different clonal trajectories are possible. The mutations can either be in completely separate cells (distinct clones) or they can be in the same cells (sub-clones). For distinct clones, different growth rates and clonal behavior would be expected, while sub clones should show similar growth rates. In this hypothetical example, individual one likely shows distinct clonal behavior while individual two likely shows sub-clone behavior. b) Bar plot showing the number of individuals categorized into the sub-clone and distinct clone groups. c) Plot showing the CHIP driver genes for each individual with two CHIP driver mutations. Each vertical line on the x-axis represents one individual and each point represents a CHIP clone. If the individual has both mutations on the same driver gene, the points are overlapping and the resulting color shows the clonal behavior.

The co-occurrence of driver genes was significantly different than what would be expected by random chance while controlling for driver gene prevalence in the cohort (X^2^= 322.03, p= 8.73 x 10^-11^). *PPM1D* and *DNMT3A* mutations co-occurred more frequently than expected (Expected: 0.9/26, Observed: 4/26, p=1), as did *DNMT3A* and *TET2* mutations (Expected: 2.8/26, Observed: 8/26, p=0.16), but this difference was not statistically significant.

### No Association of Demographic Factors with CHIP Clonal expansion

Previous studies have suggested that demographic factors may contribute to CHIP prevalence, expansion rate and risk of subsequent disease. In particular, there has been much speculation about the potential role of obesity or other metabolic factors and CHIP expansion. However, we did not detect any significant effect of age (p=0.63), gender (p=0.68), self-reported race (p=0.94), ethnicity (p=0.77), BMI (p=0.37), or height (p=0.98) on clonal expansion rate across individuals in this cohort while controlling for driver gene identity (**Supplemental Table 3**).

## Discussion

Here we present a highly scalable and cost-efficient CHIP detection assay optimized for the detection of CHIP in large-scale human cohorts. As a demonstration of the utility of this targeted assay we profiled a set of multi-timepoint blood samples from the Vanderbilt University BioVU biobank. Insights from this cohort showed the influence of driver gene, mutation type, mutation quantity, and demographic information on clonal expansion rate over time.

CHIP can be detected using whole genome/exome sequencing, but these methods are of much higher cost and complexity that is not necessary to understand CHIP clonal dynamics. Our targeted gene panel approach is low cost and is therefore a much more practical approach for large-scale CHIP studies. Currently the Vanderbilt VANTAGE core laboratory performs this assay on a cost recovery fee-for-service basis for ∼$8.40/sample which is ∼30-50-fold less expensive than alternative technologies. Approximately 80,000 samples have been run in the past 12 months for investigators at 30 institutions across the country and incorporated into multiple published reports.^14,23–25^ The key innovations in our assay include miniaturizing and identifying the optimal stoichiometry of specific DNA library preparation and capture components and identifying an 44kb DNA target region capable of detecting >95% of CHIP in the population. Our assay can accurately identify CHIP mutations and quantify them effectively as they persist over time. Due to the deep sequencing coverage of the assay, we were able to detect clonal fractions of CHIP mutations below the typical 2% VAF cutoff. Although individuals with CHIP clones of this size are less likely to progress to disease, previous studies have shown that there is still an increased risk of blood cancer and heart failure.^26,27^ Further work is needed to fully characterize the clinical risk of these small clones, and will be made possible by assays like this CHIP assay with the power to detect small clones beneath the limit of detection of exome/genome sequencing.

The application of the CHIP assay to the Vanderbilt BioVU participants revealed interesting observations about clonal behavior over time. In addition to our report, there have been three other recent papers describing CHIP expansion rate in cohorts with multi-timepoint blood samples.^19,28,29^ Our results, are highly concordant with previous studies, showing that CHIP mutations are enriched in older individuals and that *DNMT3A* and *TET2* are the most common CHIP driver genes.^9^ In our cohort, *JAK2* and *PPM1D* mutations are the next two most common mutations followed by *ASXL1*. *JAK2* mutations are enriched in our set of BioVU samples because *JAK2 p.V617F* is directly genotyped on the BioVU MEGA array and we intentionally included samples we predicted had CHIP. Upon chart review, 56% of individuals with *PPM1D* mutations underwent chemotherapy, so the overrepresentation of PPM1D is likely related to the fact that BioVU is a tertiary hospital-based cohort.

We observe that most individuals with CHIP have a single driver mutation, but that there are individuals in the cohort with multiple driver mutations. Concordant with previous work, we observed that *TET2* surpasses *DNMT3A* as the most common driver gene for individuals 75 and older.^28^

Understanding of the factors that contribute to clonal expansion is important because faster growing clones have been associated with greater risk of disease.^18,26,30^ Like other prior work, we find that the driver gene plays a large role in determining the rate of clonal growth.^28^ However, we also find that the driver gene mutation is not deterministic and that there is a significant level of individual-level heterogeneity. When we focused on specific driver mutations within the same gene in *DNMT3A,*we did not observe any significant differences in average growth rate between the two hotspot mutations, R882C and R882H. We also did not observe a significant difference between loss of function and missense mutations, although the sample size for loss of function group (n=6) precludes robust inference. Future efforts with larger sample sizes or a meta-analysis across existing published data may power further analyses at the individual mutation level.

Here we add to the recent surprising observations in comparable cohorts,^19,28^ that CHIP clones do not universally grow over time, but can also remain stagnant or shrink. While most clones grew, ∼25% of clones displayed shrinkage behavior, and shrinking clones persisted even when looking at individuals with only one CHIP clone. It is possible that these clones shrink due to the presence of an additional CHIP/non-CHIP clone that is more fit.^29^ Within individuals with two driver mutations, *DNMT3A* clones were more likely than other driver genes to remain stagnant or shrink over time, which is consistent with their slower growth rate on average. The vast majority of individuals with two CHIP clones had at least one clone that was expanding. Further studies are needed to better characterize differences in clonal behavior trajectories.

When individuals have two CHIP driver mutations, it is possible that either those mutations occur in distinct sub-populations of cells or that they co-occur in sub-clonal populations. Single timepoint data does not allow for any distinction between these possibilities because the VAF of the mutations would be the same in both cases. However, multi-timepoint cohorts provide additional information about clone behavior over time that can provide insight into which of these scenarios is most likely. In our cohort, we observed a nearly 50-50 split in distinct vs sub-clone individuals, indicating that both possibilities occur in individuals with CHIP. We did not observe a significant difference in average growth rate of the fastest growing clone between these two groups. The distribution of driver genes was not random, and combinations of *DNMT3A* & *PPM1D* and *DNMT3A* & *TET2* mutations were particularly enriched, even when controlling for their overall prevalence in the cohort. There may be underlying biology that underlies the co-occurrence of these mutations, but this could also be an artifact of the hospital-based cohort. Future studies are required to resolve whether these two scenarios confer differential disease risk.

While age and BMI have been previously associated with increased CHIP prevalence,^31^ and metabolic factors have been suggested to increase CHIP expansion rate,^32,33^ in this study, we did not detect any participant characteristics that had a significant effect on clonal growth rate. This is concordant with previous work that also did not detect any associations with clonal expansion rate and race, ethnicity, or BMI.^19^ Larger studies are needed to identify the genetic and environmental influences on the rate of clonal expansion.

Our work has limitations that are important to consider. First, although our study is one of the largest multi-timepoint longitudinal studies of CHIP, the cohort sample size is small in absolute terms, meaning that individual-level outliers have potential to influence the general conclusions of the study. Second, a range of time between blood draws is represented in our study from 6 months to 12 years. However, the process of calculating clonal growth rate (Methods) accounts for both starting VAF and time between blood draws. Third, the CHIP assay was designed to capture >95% of CHIP mutations, meaning that it is unable to capture the entire range of CHIP mutations and we do not detect larger structural changes (such as large-scale chromosomal deletions or loss of heterozygosity) that can also cause clonal expansion. Efforts are underway to develop a second generation assay to detect larger scale alterations. Fourth, although the assay is able to achieve deep sequencing depth, growth/shrinkage can still be affected by biological factors, such as the relative proportion of myeloid cells at the time of blood draw.

In summary, we developed a cost-effective and accurate tool to call CHIP mutations in large-scale human cohorts. The application of this tool to a multi-timepoint CHIP cohort in the Vanderbilt BioVU biobank revealed important observations, such as the influence of driver gene on clonal growth and the various clonal behavior trajectories observed across driver genes. Use of this assay in >80,000 samples for investigators around the world has already enabled large-scale studies to characterize the influence of CHIP on human health. We envision that this highly cost effective tool will be of particular utility for the large scale screening that will be required for identifying participants for future CHIP clinical trials.

## Data Availability

All data produced in the present study are available upon reasonable request to the authors.

## Acknowledgements

A.J.S. received financial support from the US National Institutes of Health (NIH) under a Ruth L. Kirschstein National Research Service Award F30DK127699 from the NIDDK and T32GM007347 from the NIGMS. MRS is a Leukemia and Lymphoma Clinical Scholar, and receives funding from the Biff Ruttenberg Foundation, Adventure Alle Fund, Beverly and George Rawlings Endowment, and the NIH 1R01 CA262287-01 and U01 OH012271-01. AGB is supported by NIH DP5 OD029586, a Burroughs Wellcome Fund Career Award for Medical Scientists, the E.P. Evans Foundation, RUNX1 Research Program, a Pew-Stewart Scholar for Cancer Research award, supported by the Pew Charitable Trusts and the Alexander and Margaret Stewart Trust, the Vanderbilt University Medical Center Brock Family Endowment and Young Ambassador Award. The Vanderbilt-Ingram Cancer Center is supported by a NIH P30 CA068485–19.

