## Supplemental Figures for "Cost-effective and scalable clonal hematopoiesis assay provides insight into clonal dynamics"

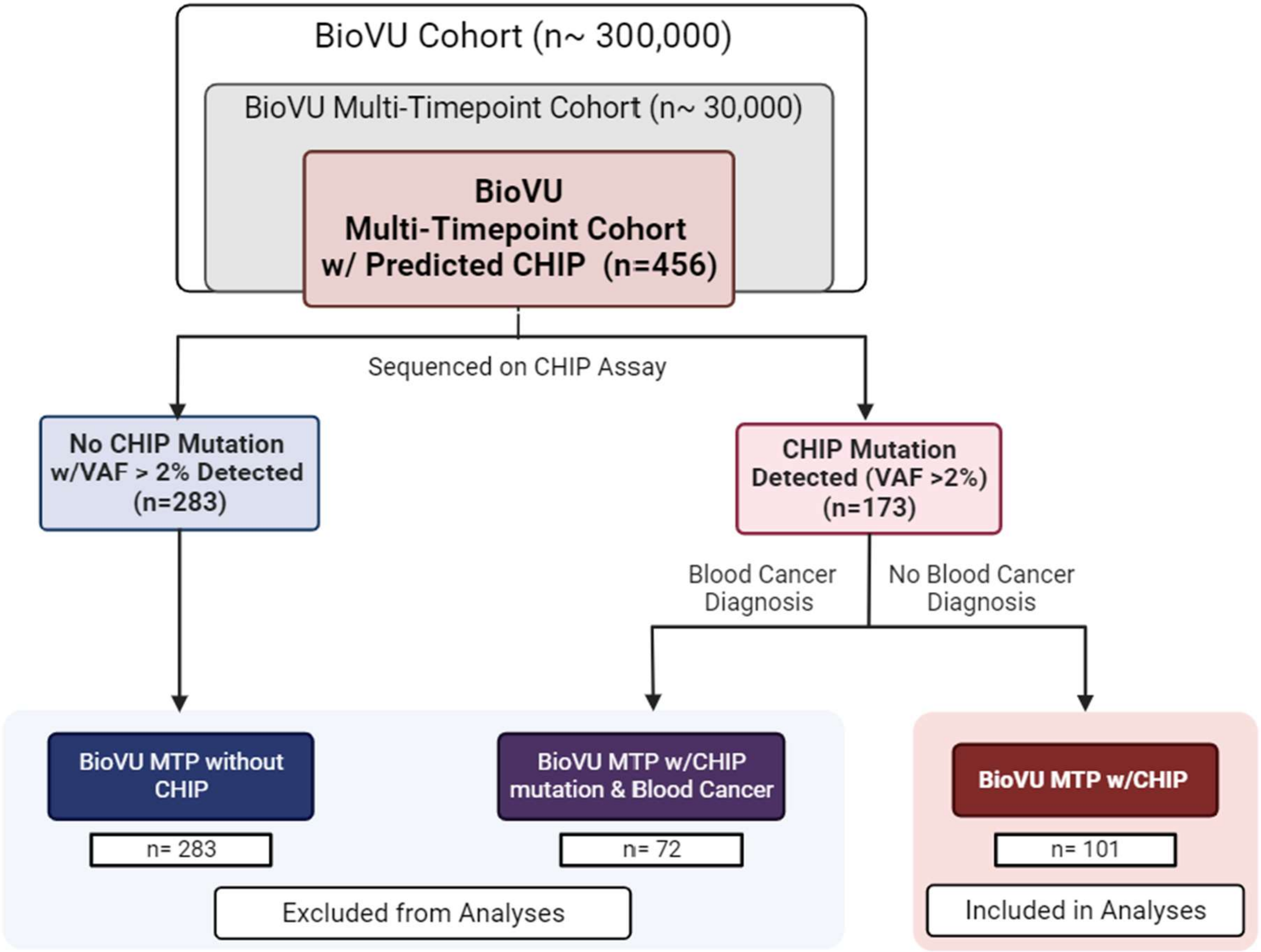


**Supplemental Figure 1: Flow chart of inclusion criteria for CHIP analyses.** A subset of individuals from the Vanderbilt BioVU cohort (n~300,000) have more than one blood sample, forming the BioVU Multi-TimePoint cohort (n~30,000). A portion of those individuals (n=456) were sequenced on the CHIP assay based on predicted CHIP. CHIP was predicted using the genotyping data from the BioVU MEGA array data. For 283 of those individuals, CHIP was not detected. For the remaining 173, patient charts were examined for the presence of blood cancer. If blood cancer was present in the record, the individual was excluded. 101 individuals remained with CHIP, and those individuals were used for further analyses.

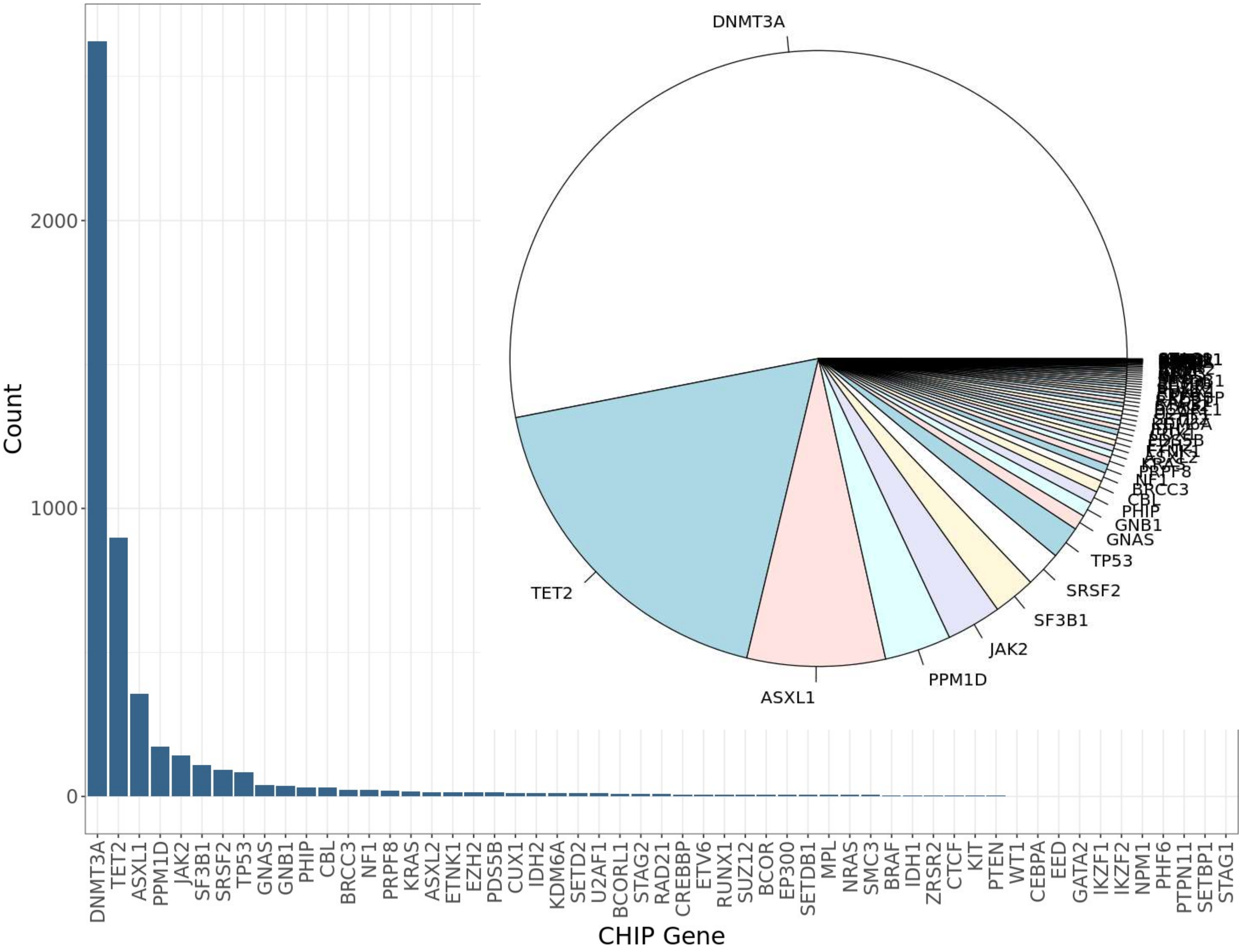


**Supplemental Figure 2: Prevalence of CHIP driver genes in TOPMed cohort (data from Bick et al, *Nature,* 2020).**[9] Bar plot and pie chart showing the distribution of CHIP driver genes across the TOPMed cohort (4,938 CHIP mutations in 4,229 individuals). >75% of mutations were in either *DNMT3A, TET2,* or *ASXL1*.

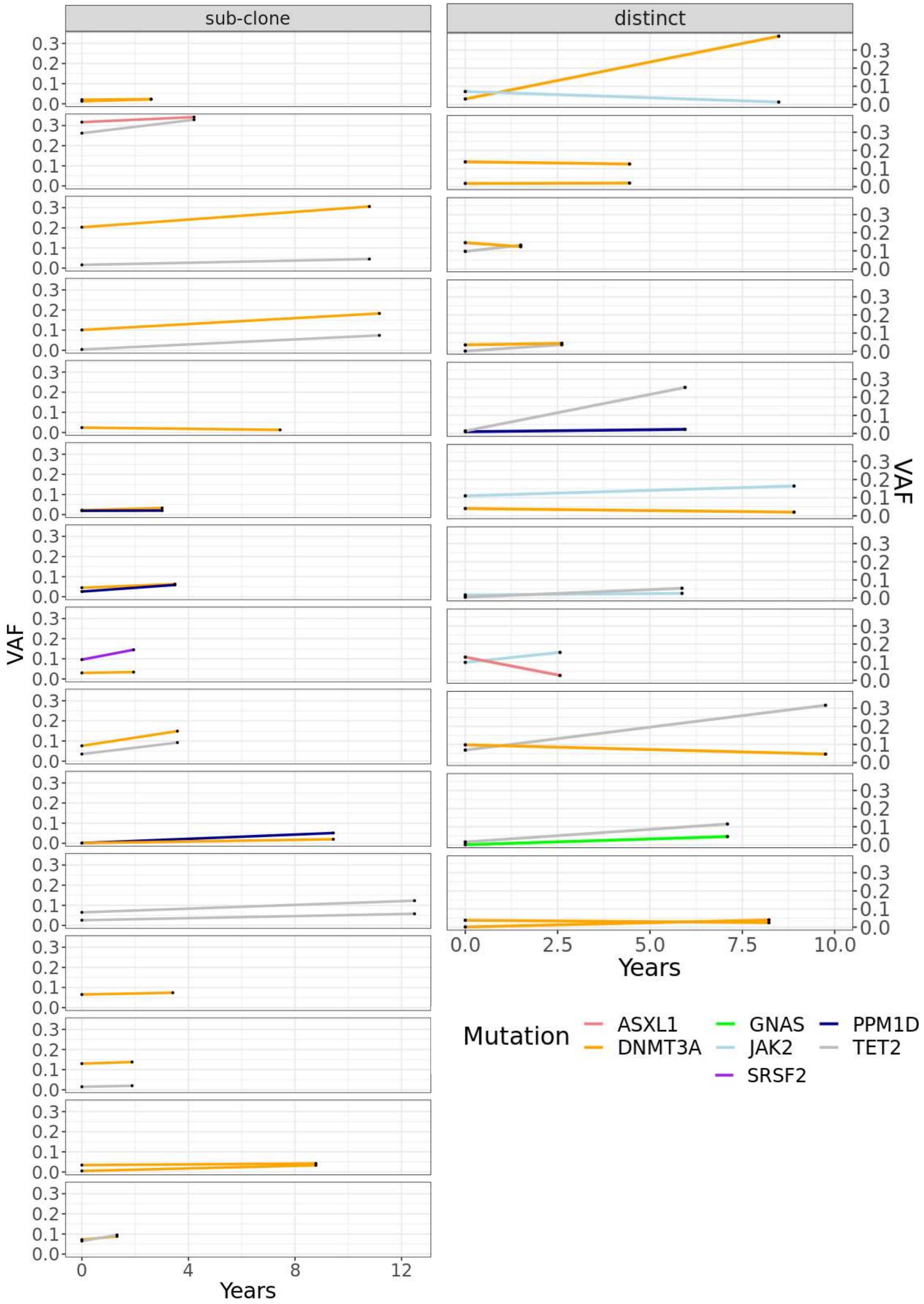


**Supplemental Figure 3: Individual clonal dynamics over time.** Plot showing the clonal behavior for each individual with two CHIP driver mutations. Each line represents one clone and each small plot represents an individual from the cohort. Facets on the left are categorized as sub-clonal while facets on the right are categorized as distinct clones.

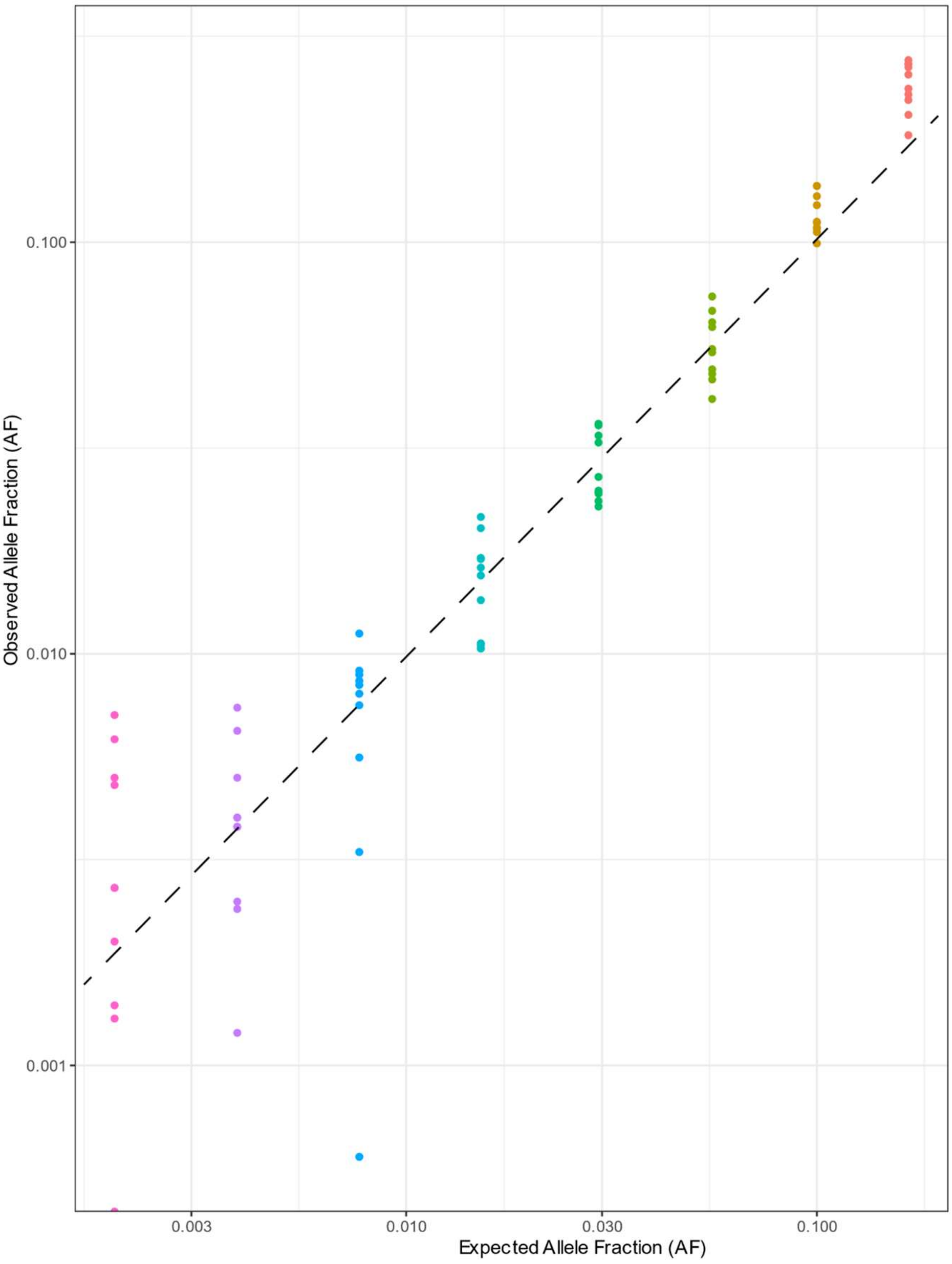


**Supplemental Figure 4: Limit of detection test for CHIP detection assay.** Results of a limiting dilution experiment where a DNA sample with known genotype was combined at serial fixed ratios with a second sample of known genotype. Each color represents a distinct sample, and the correlation line is displayed to show the correlation between observed and expected AF. Our method robustly detects variants present in >1% of DNA. Furthermore, beneath this 1% threshold, we continue to detect variants down to ~0.1% allele fraction, but with less accuracy for the estimated allele fraction.
