## Supplemental Tables for "Cost-effective and scalable clonal hematopoiesis assay provides insight into clonal dynamics"

**Supplementary Tables**

**Supplemental Table 1: Descriptive statistics on study cohort and demographic information.**

| **Cohort** | **Number of Individuals** | **Mean Age (Timepoint 1)** | **Mean Age (Timepoint 2)** | **Biological Sex** | **Self-Reported Race** |
| --- | --- | --- | --- | --- | --- |
| BioVU MTP w/Predicted CHIP | 456 | 52.5 years  (SD: 19 years) | 57 years (SD: 19 years) | 60% female | 81% Caucasian  15% African American  1% Asian |
| BioVU MTP without CHIP | 283 | 47 years (SD: 19 years) | 52 years (SD: 19 years) | 59% female | 77% Caucasian  19% African American  2% Asian |
| BioVU MTP w/CHIP mutation & Blood Cancer | 72 | 60 years (SD: 17 years) | 64 years (SD: 17 years) | 63% female | 93% Caucasian  4% African American |
| **BioVU MTP w/CHIP** | 101 | 64 years (SD: 14 years) | 69 years (SD: 14 years) | 64% female | 88% Caucasian  10% African American |

**Supplemental Table 2:** Targeted Capture regions for identifying an individual with CHIP. Genomic coordinates are provided per the hg38 reference genome.

| **Chromosome** | **Start** | **Stop** | **Gene** |
| --- | --- | --- | --- |
| chr1 | 1806469 | 1806543 | *GNB1* |
| chr1 | 1815750 | 1815867 | *GNB1* |
| chr1 | 43349257 | 43349363 | *MPL* |
| chr1 | 114713799 | 114713978 | *NRAS* |
| chr1 | 114716047 | 114716162 | *NRAS* |
| chr2 | 25749689 | 25750420 | *ASXL2* |
| chr2 | 25753532 | 25753640 | *ASXL2* |
| chr2 | 25246614 | 25246781 | *DNMT3A* |
| chr2 | 25313907 | 25313989 | *DNMT3A* |
| chr2 | 25234273 | 25234425 | *DNMT3A* |
| chr2 | 25235701 | 25235830 | *DNMT3A* |
| chr2 | 25236930 | 25237010 | *DNMT3A* |
| chr2 | 25239124 | 25239220 | *DNMT3A* |
| chr2 | 25239484 | 25239518 | *DNMT3A* |
| chr2 | 25240296 | 25240455 | *DNMT3A* |
| chr2 | 25240634 | 25240735 | *DNMT3A* |
| chr2 | 25241556 | 25241712 | *DNMT3A* |
| chr2 | 25243892 | 25243987 | *DNMT3A* |
| chr2 | 25244149 | 25244343 | *DNMT3A* |
| chr2 | 25244534 | 25244657 | *DNMT3A* |
| chr2 | 25245247 | 25245337 | *DNMT3A* |
| chr2 | 25246014 | 25246069 | *DNMT3A* |
| chr2 | 25246154 | 25246314 | *DNMT3A* |
| chr2 | 25247045 | 25247163 | *DNMT3A* |
| chr2 | 25247585 | 25247754 | *DNMT3A* |
| chr2 | 25248031 | 25248257 | *DNMT3A* |
| chr2 | 25249651 | 25249729 | *DNMT3A* |
| chr2 | 25251906 | 25252099 | *DNMT3A* |
| chr2 | 25252188 | 25252202 | *DNMT3A* |
| chr2 | 25274935 | 25275092 | *DNMT3A* |
| chr2 | 25275494 | 25275548 | *DNMT3A* |
| chr2 | 25282382 | 25282716 | *DNMT3A* |
| chr2 | 25300133 | 25300248 | *DNMT3A* |
| chr2 | 208243524 | 208243601 | *IDH1* |
| chr2 | 208248358 | 208248421 | *IDH1* |
| chr2 | 197400709 | 197400941 | *SF3B1* |
| chr2 | 197401979 | 197402135 | *SF3B1* |
| chr2 | 197405269 | 197405477 | *SF3B1* |
| chr2 | 197416735 | 197416916 | *SF3B1* |
| chr2 | 197400049 | 197400171 | *SF3B1* |
| chr2 | 197400246 | 197400439 | *SF3B1* |
| chr2 | 197401394 | 197401530 | *SF3B1* |
| chr2 | 197401736 | 197401893 | *SF3B1* |
| chr2 | 197402550 | 197402831 | *SF3B1* |
| chr2 | 197402943 | 197403040 | *SF3B1* |
| chr2 | 197403579 | 197403769 | *SF3B1* |
| chr2 | 197405070 | 197405182 | *SF3B1* |
| chr2 | 197407992 | 197408124 | *SF3B1* |
| chr2 | 197408363 | 197408586 | *SF3B1* |
| chr2 | 197409764 | 197410012 | *SF3B1* |
| chr4 | 54727217 | 54727324 | *KIT* |
| chr4 | 54733069 | 54733192 | *KIT* |
| chr4 | 54727415 | 54727542 | *KIT* |
| chr4 | 54727822 | 54727927 | *KIT* |
| chr4 | 54728010 | 54728121 | *KIT* |
| chr4 | 54729334 | 54729485 | *KIT* |
| chr4 | 54731327 | 54731419 | *KIT* |
| chr4 | 54731870 | 54731998 | *KIT* |
| chr4 | 54736497 | 54736609 | *KIT* |
| chr4 | 105233891 | 105237445 | *TET2* |
| chr4 | 105243564 | 105243783 | *TET2* |
| chr4 | 105269604 | 105269752 | *TET2* |
| chr4 | 105275042 | 105276524 | *TET2* |
| chr4 | 105241333 | 105241438 | *TET2* |
| chr4 | 105242828 | 105242932 | *TET2* |
| chr4 | 105259613 | 105259774 | *TET2* |
| chr4 | 105261753 | 105261853 | *TET2* |
| chr4 | 105272558 | 105272923 | *TET2* |
| chr9 | 5076681 | 5076701 | *JAK2* |
| chr9 | 5073683 | 5073800 | *JAK2* |
| chr11 | 119278160 | 119278302 | *CBL* |
| chr11 | 119278504 | 119278718 | *CBL* |
| chr12 | 22671265 | 22671359 | *ETNK1* |
| chr12 | 25225612 | 25225772 | *KRAS* |
| chr12 | 25227220 | 25227424 | *KRAS* |
| chr12 | 25245270 | 25245384 | *KRAS* |
| chr15 | 90088655 | 90088758 | *IDH2* |
| chr17 | 60656593 | 60656846 | *PPM1D* |
| chr17 | 60662989 | 60663557 | *PPM1D* |
| chr17 | 7669603 | 7669695 | *TP53* |
| chr17 | 7670603 | 7670720 | *TP53* |
| chr17 | 7673213 | 7673271 | *TP53* |
| chr17 | 7673301 | 7673344 | *TP53* |
| chr17 | 7673529 | 7673613 | *TP53* |
| chr17 | 7673695 | 7673842 | *TP53* |
| chr17 | 7674175 | 7674295 | *TP53* |
| chr17 | 7674814 | 7674976 | *TP53* |
| chr17 | 7675047 | 7675243 | *TP53* |
| chr17 | 7675988 | 7676277 | *TP53* |
| chr17 | 7676376 | 7676408 | *TP53* |
| chr17 | 7676515 | 7676627 | *TP53* |
| chr18 | 44951903 | 44952002 | *SETBP1* |
| chr20 | 32358770 | 32358837 | *ASXL1* |
| chr20 | 32359741 | 32359796 | *ASXL1* |
| chr20 | 32366378 | 32366472 | *ASXL1* |
| chr20 | 32369006 | 32369128 | *ASXL1* |
| chr20 | 32428122 | 32428253 | *ASXL1* |
| chr20 | 32428319 | 32428427 | *ASXL1* |
| chr20 | 32429332 | 32429436 | *ASXL1* |
| chr20 | 32429895 | 32430058 | *ASXL1* |
| chr20 | 32431315 | 32431489 | *ASXL1* |
| chr20 | 32431577 | 32431684 | *ASXL1* |
| chr20 | 32432874 | 32432990 | *ASXL1* |
| chr20 | 32433278 | 32433922 | *ASXL1* |
| chr20 | 32434426 | 32437343 | *ASXL1* |
| chr20 | 58910677 | 58910834 | *GNAS* |
| chr20 | 58909344 | 58909428 | *GNAS* |
| chr20 | 58909515 | 58909584 | *GNAS* |
| chr20 | 58909678 | 58909809 | *GNAS* |
| chr20 | 58909945 | 58910086 | *GNAS* |
| chr20 | 58910328 | 58910406 | *GNAS* |
| chr21 | 43094649 | 43094793 | *U2AF1* |
| chr21 | 43093096 | 43093254 | *U2AF1* |
| chr21 | 43094461 | 43094568 | *U2AF1* |
| chr21 | 43095432 | 43095541 | *U2AF1* |
| chr21 | 43095688 | 43095748 | *U2AF1* |
| chr21 | 43100447 | 43100524 | *U2AF1* |
| chr21 | 43101274 | 43101437 | *U2AF1* |
| chr21 | 43104309 | 43104407 | *U2AF1* |
| chr21 | 43107445 | 43107499 | *U2AF1* |
| chrX | 155071527 | 155071650 | *BRCC3* |
| chrX | 155072326 | 155072343 | *BRCC3* |

**Supplemental Table 3: Regression analysis of clonal growth rate with participant characteristics.**

| **Participant Characteristic** | **Beta Value** | **95% CI** | **P-Value** |
| --- | --- | --- | --- |
| Age at Timepoint 1 | -0.003 | [-0.02, 0.01] | 0.63 |
| Gender | -0.09 | [-0.53, 0.34] | 0.68 |
| Self-reported Race | -0.09 | [-2.37, 2.19] | 0.94 |
| Ethnicity | 0.36 | [-2.04, 2.76] | 0.77 |
| BMI | 0.02 | [-0.02, 0.05] | 0.37 |
| Height | -0.0003 | [-0.02, 0.02] | 0.98 |
